# Genome-wide association study identifies a new susceptibility locus in *PLA2G4C* for Multiple System Atrophy

**DOI:** 10.1101/2023.05.02.23289328

**Authors:** Yasuo Nakahara, Jun Mitsui, Hidetoshi Date, Kristine Joyce Porto, Yasuhiro Hayashi, Atsushi Yamashita, Yoshio Kusakabe, Takashi Matsukawa, Hiroyuki Ishiura, Tsutomu Yasuda, Atsushi Iwata, Jun Goto, Yaeko Ichikawa, Yoshio Momose, Yuji Takahashi, Tatsushi Toda, Rikifumi Ohta, Jun Yoshimura, Shinichi Morishita, Emil K Gustavsson, Darren Christy, Melissa Maczis, Matthew J. Farrer, Han-Joon Kim, Sung-Sup Park, Beomseok Jeon, Jin Zhang, Weihong Gu, Sonja W. Scholz, Andrew B. Singleton, Henry Houlden, Ichiro Yabe, Hidenao Sasaki, Masaaki Matsushima, Hiroshi Takashima, Akio Kikuchi, Masashi Aoki, Kenju Hara, Akiyoshi Kakita, Mitsunori Yamada, Hitoshi Takahashi, Osamu Onodera, Masatoyo Nishizawa, Hirohisa Watanabe, Mizuki Ito, Gen Sobue, Kinya Ishikawa, Hidehiro Mizusawa, Kazuaki Kanai, Satoshi Kuwabara, Kimihito Arai, Shigeru Koyano, Yoshiyuki Kuroiwa, Kazuko Hasegawa, Tatsuhiko Yuasa, Kenichi Yasui, Kenji Nakashima, Hijiri Ito, Yuishin Izumi, Ryuji Kaji, Takeo Kato, Susumu Kusunoki, Yasushi Osaki, Masahiro Horiuchi, Ken Yamamoto, Mihoko Shimada, Taku Miyagawa, Yosuke Kawai, Nao Nishida, Katsushi Tokunaga, Alexandra Dürr, Alexis Brice, Alessandro Filla, Thomas Klockgether, Ullrich Wüllner, Caroline M. Tanner, Walter A. Kukull, Virginia M.-Y. Lee, Eliezer Masliah, Phillip A. Low, Paola Sandroni, Laurie Ozelius, Tatiana Foroud, Shoji Tsuji

## Abstract

To elucidate the molecular basis of multiple system atrophy (MSA), a neurodegenerative disease, we conducted a genome-wide association study (GWAS) in a Japanese MSA case/control series followed by replication studies in Japanese, Korean, Chinese, European and North American samples. In the GWAS stage rs2303744 on chromosome 19 showed a suggestive association (*P* = 6.5 × 10^−7^) that was replicated in additional Japanese samples (*P* = 2.9 × 10^−6^. OR = 1.58; 95% confidence interval, 1.30 to 1.91), and then confirmed as highly significant in a meta-analysis of East Asian population data (*P* = 5.0 × 10^-15^. Odds ratio= 1.49; 95% CI 1.35 to 1.72). The association of rs2303744 with MSA remained significant in combined European/North American samples (*P* =0.023. Odds ratio=1.14; 95% CI 1.02 to 1.28) despite allele frequencies being quite different between these populations. rs2303744 leads to an amino acid substitution in *PLA2G4C* that encodes the cPLA2γ lysophospholipase/transacylase. The cPLA2γ-Ile143 isoform encoded by the MSA risk allele has significantly decreased transacylase activity compared with the alternate cPLA2γ-Val143 isoform that may perturb membrane phospholipids and α-synuclein biology.

---

Multiple system atrophy (MSA) is a neurodegenerative disease characterized by autonomic failure with various combinations of parkinsonism and cerebellar ataxia ^1, 2^. The cardinal neuropathological hallmark of MSA is the presence of argyrophilic glial cytoplasmic inclusions (GCIs) of which α–synuclein is a major component^1, 2^. MSA is classified into two subtypes, MSA-C (characterized predominantly by cerebellar ataxia) and MSA-P (characterized predominantly by parkinsonism)^3^. The relative frequencies of MSA-P compared to MSA-C in Europe and North America are reported as 63% and 60% of cases, while MSA-C is reported in 65–67% of cases in Japan^4–8^. Although MSA is largely considered to have no heritable component^9^, several multiplex MSA families with confirmed neuropathological diagnoses have been reported^10–12^.

To elucidate the molecular basis of MSA, association studies focusing on candidate genes^13–25^ including *SNCA*^20–22, 25^, *LRRK2*^24^, and *GBA*^15^ have been conducted. A genome-wide association study (GWAS) on 918 MSA patients of European ancestry and 3,864 controls revealed several potentially interesting gene loci^26^. Focusing on multiplex MSA families, we identified homozygous or compound heterozygous *COQ2* mutations in two multi-incident MSA families and implicated functionally impaired *COQ2* variants in sporadic MSA^27^. *COQ2* p.Val393Ala is a common polymorphism in East Asian populations, and meta-analyses have demonstrated significant associations of COQ2 p.Val393Ala with sporadic MSA^28, 29^ . It should be noted that *COQ2* p.Val393Ala was not informative in European and North American populations as it is monomorphic.

To further elucidate the molecular basis of MSA, we conducted a GWAS of MSA in Japanese samples. We replicated the genome-wide association in East Asian samples to directly identify a coding variant in a new susceptibility gene, *PLA2G4C,* that affects the transacylation activity of the enzyme encoded.

## RESULTS

### Association of rs2303744 located in exon 5 of *PLA2G4C* with MSA

We conducted GWAS with a total of 382 cases and 385 controls originating from Japan (**Table 1**). Genome-wide significance was set at *P* < 1.09 × 10^-7^ using a stringent Bonferroni’s correction to take into account the 456,818 informative SNPs examined. Quartile-quartile (QQ) plots of observed and expected *P* value distributions showed no systematic bias or unrecognized population structure (λ_GC_, 0.995) (**Supplementary Fig. 1**). **Fig. 1A** provides a Manhattan plot of the single-point association data for genotypes obtained. Although no loci with genome-wide significance (*P*<1.1 × 10^-7^) were observed, rs2303744 on chromosome 19 (GRCh38 NC_000019.10:g.48099691T>C (plus (+) strand)) showed a suggestive association with *P* = 6.5 × 10^-7^ (**Fig. 1**). Note *PLA2G4C* is in the reverse orientation and rs2303744 in exon 5 denotes an amino acid substitution corresponding to Ile [<u>A</u>TC] > Val [<u>G</u>TC] at codon 143 (NM_003706). rs2303744 is the most highly associated SNP in the locus albeit with a block of neighboring SNPs spanning the adjacent gene, *LIG1*, upstream of the *PLA2G4C* promoter. *LIG1* variants are in linkage disequilibrium and less associated given relatively high rates of population recombination within *PLA2G4C,* flanking rs2303744, that compromises its pairwise correlation with neighboring variants (**Fig. 1B**). Since the major allele of rs2303744 is G in East Asian populations, odds ratios were subsequently calculated based on the G allele as the reference.

**Figure 1.**
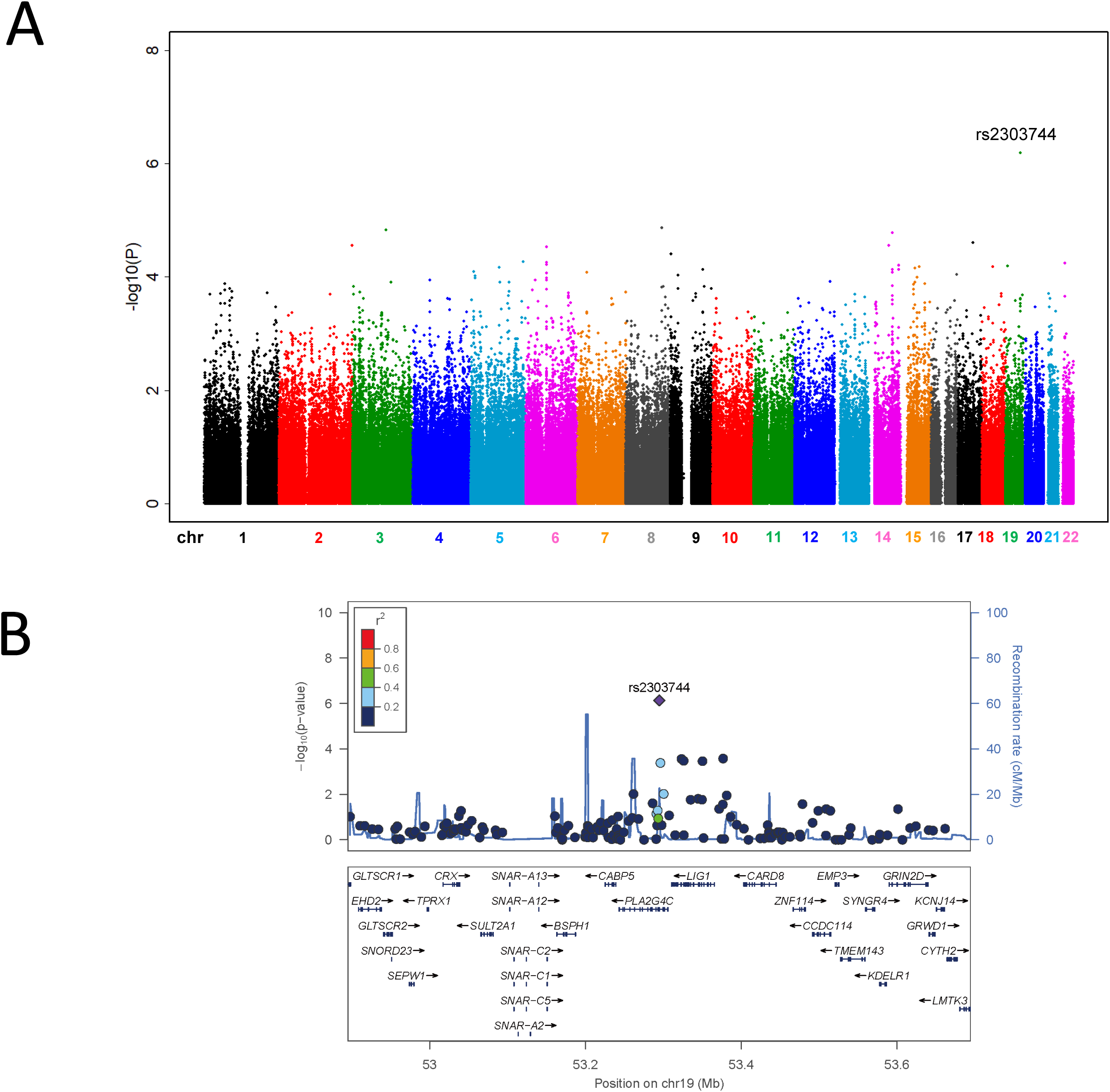
A. Genome-wide Manhattan plots for MSA (Japanese GWAS series). Plot shows the individual *P* values of SNPs calculated using a χ2 test for genotype counts highlighting the candidate SNP, rs2303744, with marginal significance (*P* = 6.5 × 10^−7^) on chromosome 19. **B.** Regional association plot of rs2303744 and its immediate locus in the Japanese GWAS series. Chromosomal position is indicated on the x axis, and the −log10 P value for each SNP is indicated on the y axis with the names and location of genes shown below. The colors of the other variants indicate their r^2^ with rs2303744.

**Table 1.**
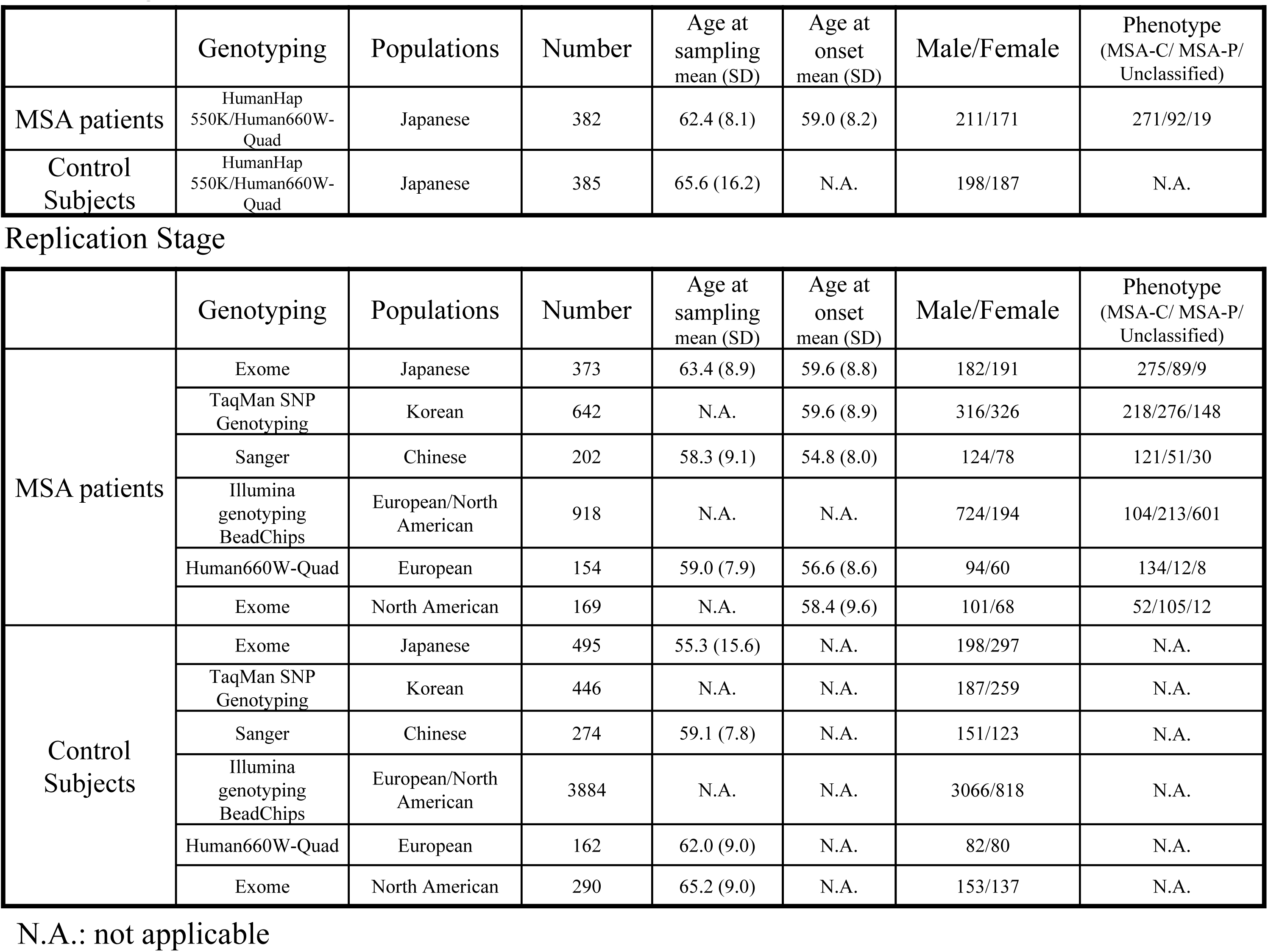
demographic data of MSA cases and controls.

|  | Genotyping | Populations | Number | Age at sampling mean (SD) | Age at onset mean (SD) | Male/Female | Phenotype (MSA-C/ MSA-P/ Unclassified) |
| --- | --- | --- | --- | --- | --- | --- | --- |
| MSA patients | HumanHap 550K/Human660W-Quad | Japanese | 382 | 62.4 (8.1) | 59.0 (8.2) | 211/171 | 271/92/19 |
| Control Subjects | HumanHap 550K/Human660W-Quad | Japanese | 385 | 65.6 (16.2) | N.A. | 198/187 | N.A. |

**Replication Stage**
|  | Genotyping | Populations | Number | Age at sampling mean (SD) | Age at onset mean (SD) | Male/Female | Phenotype (MSA-C/ MSA-P/ Unclassified) |
| --- | --- | --- | --- | --- | --- | --- | --- |
| MSA patients | Exome | Japanese | 373 | 63.4 (8.9) | 59.6 (8.8) | 182/191 | 275/89/9 |
|  | TaqMan SNP Genotyping | Korean | 642 | N.A. | 59.6 (8.9) | 316/326 | 218/276/148 |
|  | Sanger | Chinese | 202 | 58.3 (9.1) | 54.8 (8.0) | 124/78 | 121/51/30 |
|  | Illumina genotyping BeadChips | European/North American | 918 | N.A. | N.A. | 724/194 | 104/213/601 |
|  | Human660W-Quad | European | 154 | 59.0 (7.9) | 56.6 (8.6) | 94/60 | 134/12/8 |
|  | Exome | North American | 169 | N.A. | 58.4 (9.6) | 101/68 | 52/105/12 |
| Control Subjects | Exome | Japanese | 495 | 55.3 (15.6) | N.A. | 198/297 | N.A. |
|  | TaqMan SNP Genotyping | Korean | 446 | N.A. | N.A. | 187/259 | N.A. |
|  | Sanger | Chinese | 274 | 59.1 (7.8) | N.A. | 151/123 | N.A. |
|  | Illumina genotyping BeadChips | European/North American | 3884 | N.A. | N.A. | 3066/818 | N.A. |
|  | Human660W-Quad | European | 162 | 62.0 (9.0) | N.A. | 82/80 | N.A. |
|  | Exome | North American | 290 | 65.2 (9.0) | N.A. | 153/137 | N.A. |

N.A.: not applicable

We replicated the chromosome 19 (rs2303744) association in second independent Japanese series (*P* = 2.9 × 10^-6^. OR = 1.58; 95% CI, 1.30 to 1.91), that combined gave a highly significant result, *P*=2.1 x 10^-11^. OR= 1.61; 95% CI, 1.40 to 1.85 (**Table 2**). The 38K JPN (Integrative Japanese Variation Database, Tohoku University Tohoku Medical Megabank Organization (ToMMo) (https://jmorp.megabank.tohoku.ac.jp/) control data provides yet more assurance (*P* = 2.2 × 10^-16^. OR = 1.62; 95% CI, 1.46 to 1.80) (**Supplementary Table 1**).

**Table 2.** Allelic association test of rs2303744 (PLA2G4C) for multiple system atrophy (MSA) in Japanese, Chinese, Korean, European and North American series.

| dbSNP<br>rs ID | Gene | Risk<br>Allele | Allele | Stage | Case |  | Control |  | OR | *P*-Value |
| --- | --- | --- | --- | --- | --- | --- | --- | --- | --- | --- |
|  |  |  |  |  | A | G | A | G |  |  |
| rs2303744 | *PLA2G4C* | A | A/G | GWAS | 380 (0.497) | 384 (0.503) | 286 (0.371) | 484 (0.629) | 1.67 (1.37-2.05) | $6.5 \times 10^{-7}$ |
| | | | | Replication | 391 (0.524) | 355 (0.476) | 407 (0.411) | 583 (0.589) | 1.58 (1.30-1.91) | $2.9 \times 10^{-6}$ |
| | | | | Combined | 771 (0.511) | 739 (0.489) | 693 (0.394) | 1067 (0.606) | 1.61 (1.40-1.85) | $2.1 \times 10^{-11}$ |
| | | | | Korean | 691 (0.538) | 593 (0.462) | 409 (0.459) | 483 (0.541) | 1.38 (1.16-1.63) | $2.6 \times 10^{-4}$ |
|  |  |  |  | Chinese | 204 (0.500) | 200 (0.500) | 238 (0.434) | 310 (0.566) | 1.33 (1.03-1.72) | 0.031 |
|  |  |  |  | European/North<br>American | 1469 (0.804) | 357 (0.196) | 6087 (0.784) | 1673 (0.216) | 1.13 (1.00-1.28) | 0.059 |
|  |  |  |  | European | 250 (0.812) | 58 (0.188) | 253 (0.781) | 71 (0.219) | 1.21 (0.82-1.78) | 0.34 |
|  |  |  |  | North American | 269 (0.796) | 69 (0.204) | 447 (0.771) | 133 (0.229) | 1.16 (0.84-1.61) | 0.37 |

Odds ratio (OR) of the effect with respect to the allele A and association *P*-values are given.

Next, we sought to extend this observation beyond Japan to independent Korean, Chinese, European and North American samples. One caveat is that frequencies of the rs2303744 A allele in Japanese, Korean and Chinese controls are 0.394, 0.459 and 0.434, respectively, while those in European/North American, European or North American controls are 0.784, 0.781 and 0.771.

Significant associations were observed in Korean (*P* = 2.6 × 10^-4^. OR = 1.38; 95% CI, 1.16 to 1.63) and Chinese series (*P* = 0.031. OR = 1.33; 95% CI, 1.03 to 1.72) (**Table 2**). Meta-analysis of East Asian populations combined (Japanese, Korean and Chinese) sums the evidence for a highly significant association between rs2303744 and MSA (*P* = 5.0 × 10^-15^. OR = 1.49; 95% CI, 1.35 to 1.65). However, no association was observed between rs2303744 and MSA in any single European or North American series (**Table 2**). Nevertheless, meta-analysis of European/North American samples combined provided marginal support (*P* = 0.023. OR = 1.14; 95% CI, 1.02 to 1.28) and meta-analysis of all sample series remained highly significant (*P* = 4.9 × 10^-6^. OR = 1.35; 95% CI, 1.19 to 1.54) (**Fig. 2**).

**Figure 2.**
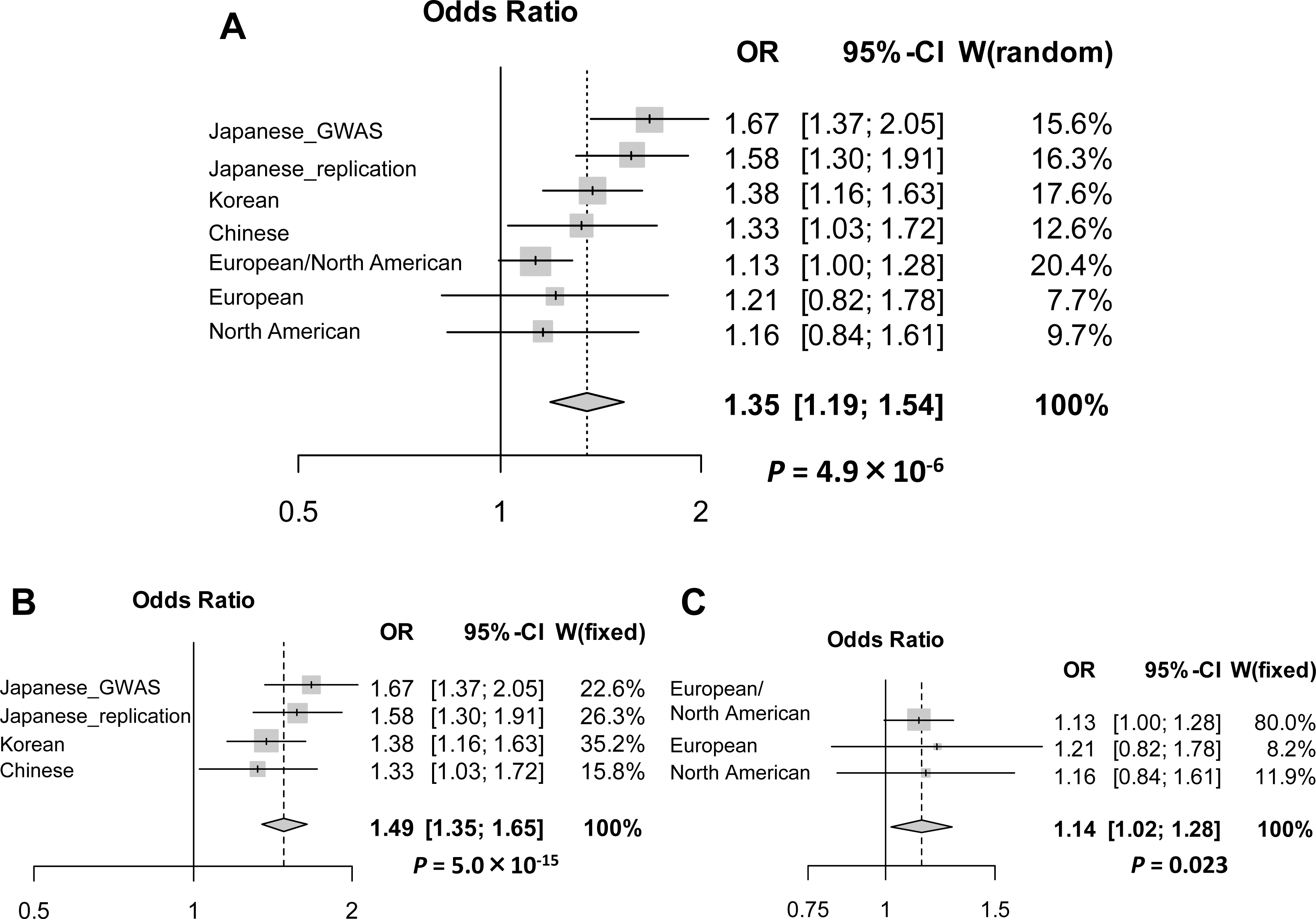
Forest plots showing the meta-analysis of rs2303744. Squares and horizontal lines represent estimated ORs and 95% CIs for individual series. The size of each square represents the statistical weight, the inverse of the variance, in each study. A diamond shows the summary OR estimate and 95% CIs for the meta-analysis. The area of diamond is proportional to the sum of the logarithm of odds ratios weighted by the inverse of the variance of each study.

Given these results we conducted multiple logistic regression analysis, adjusted for sex and population, to evaluate rs2303744 genotypic associations with MSA in East Asian samples, for which a significant association was demonstrated (*P* = 6.6 × 10^-16^. OR=1.51; 95% CI, 1.37 to 1.67). Intriguingly, multiple linear regression analysis adjusted for sex and population demonstrates a correlation between rs2303744 genotype and the age at onset of MSA, with AA genotypes showing the youngest mean ages at onset in the Japanese, Korean and Chinese samples (*P* = 0.0010) (**Fig. 3**). A meta-analysis including Japanese, Korean and Chinese populations, a recessive model demonstrated a most evidence for association of the AA genotype with MSA (*P* = 1.8 × 10^-12^. OR=1.86; 95% CI, 1.57 to 2.21). When MSA cases are subdivided into MSA-C and MSA-P cases, multiple logistic regression analysis adjusted for sex and population showed significant associations in MSA-C (*P* = 8.6 × 10^-14^. OR = 1.57; 95% CI, 1.39 to 1.77) and MSA-*P* (*P* = 7.2 × 10^-6^. OR= 1.38; 95% CI, 1.20 to 1.59) in East Asian populations (**Supplementary Table 2**).

**Figure 3.**
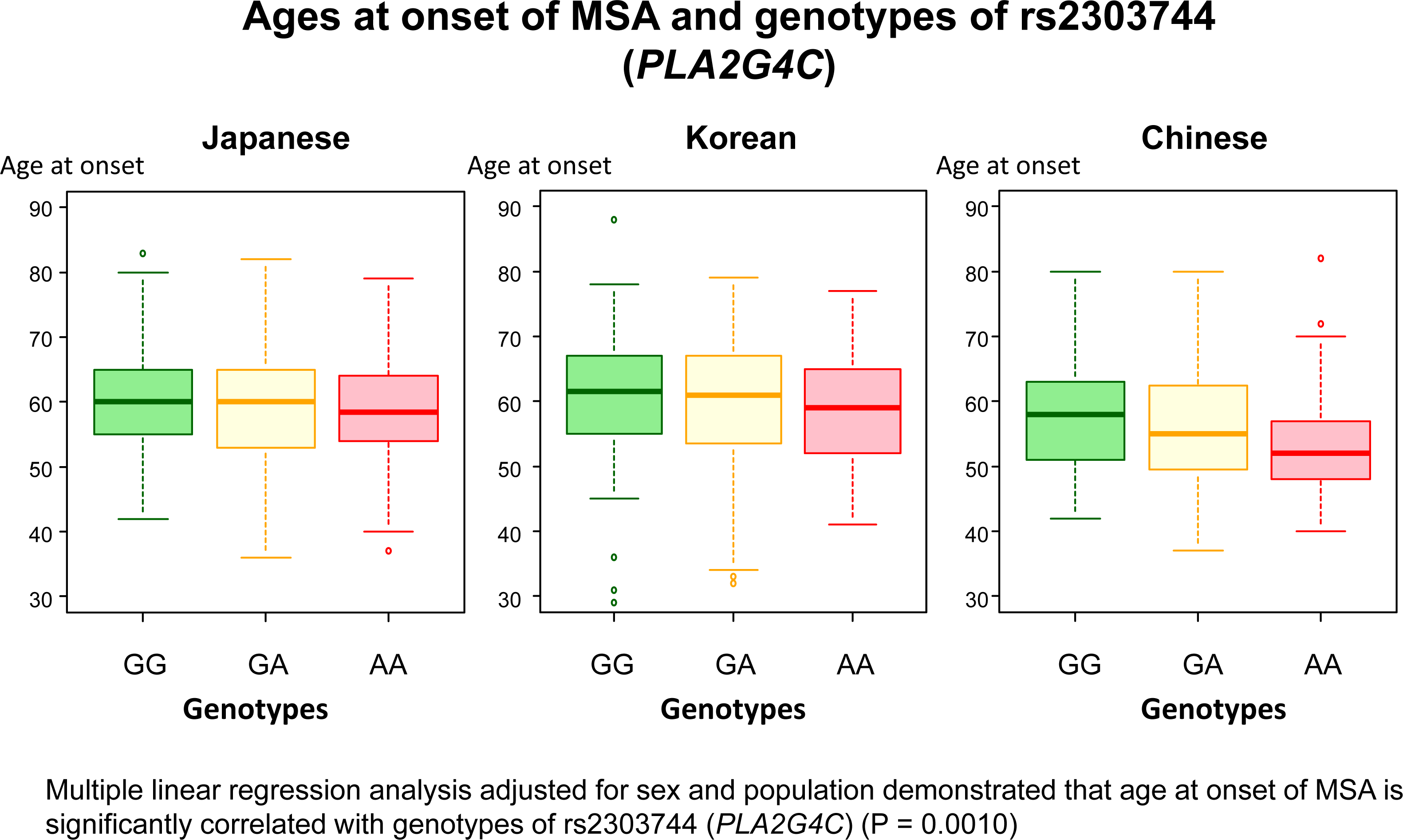
Ages at onset of MSA and genotypes of rs2303744 (PLA2G4C) In the Japanese series, mean ages at onset of MSA patients with AA, AG, and GG genotypes are 58.8, 59.2 and 60.0. In the Korean series, mean ages at onset of MSA patients with AA, AG, and GG genotypes are 58.6, 59.6 and 61.0, respectively. In the Chinese series, mean ages at onset of MSA patients with AA, AG, and GG genotypes are 53.3, 55.8 and 57.8, respectively. Multiple liner regression analysis adjusted for sex and population demonstrated that age at onset of MSA is significantly correlated with genotypes of rs2303744 (*PLA2G4C*) (*P* = 0.001).

### Functional evaluation of *PLA2G4C* with rs2303744

Functional analysis of transiently expressed cPLA2γ, the gene product of *PLA2G4C*, was conducted to examine the effect of the rs2303744 SNP [*PLA2G4C* c.427A>G (cPLA2γ p.Ile143Val) (NM_001159323.1; NP_001152795.1)]. Recombinant proteins for alternate alleles were transiently expressed in HEK293 cells and lysophospholipase/transacylation activities were examined. Western blotting revealed the expression of each allele was not significantly altered (**Fig. 4A**). When the substrate [^14^C]lysophosphatidylcholine (LPC, 1-[^14^C]oleoyl-glycerophosphocholine (GPC)) was incubated with protein extract from FLAG-cPLA2γ-transfected HEK293 cells, both [^14^C]FFA ([^14^C]oleic acid) and [^14^C]PC(1,2-diacyl-GPC) reaction products were observed (**Fig. 4B, C**).

**Figure 4.**
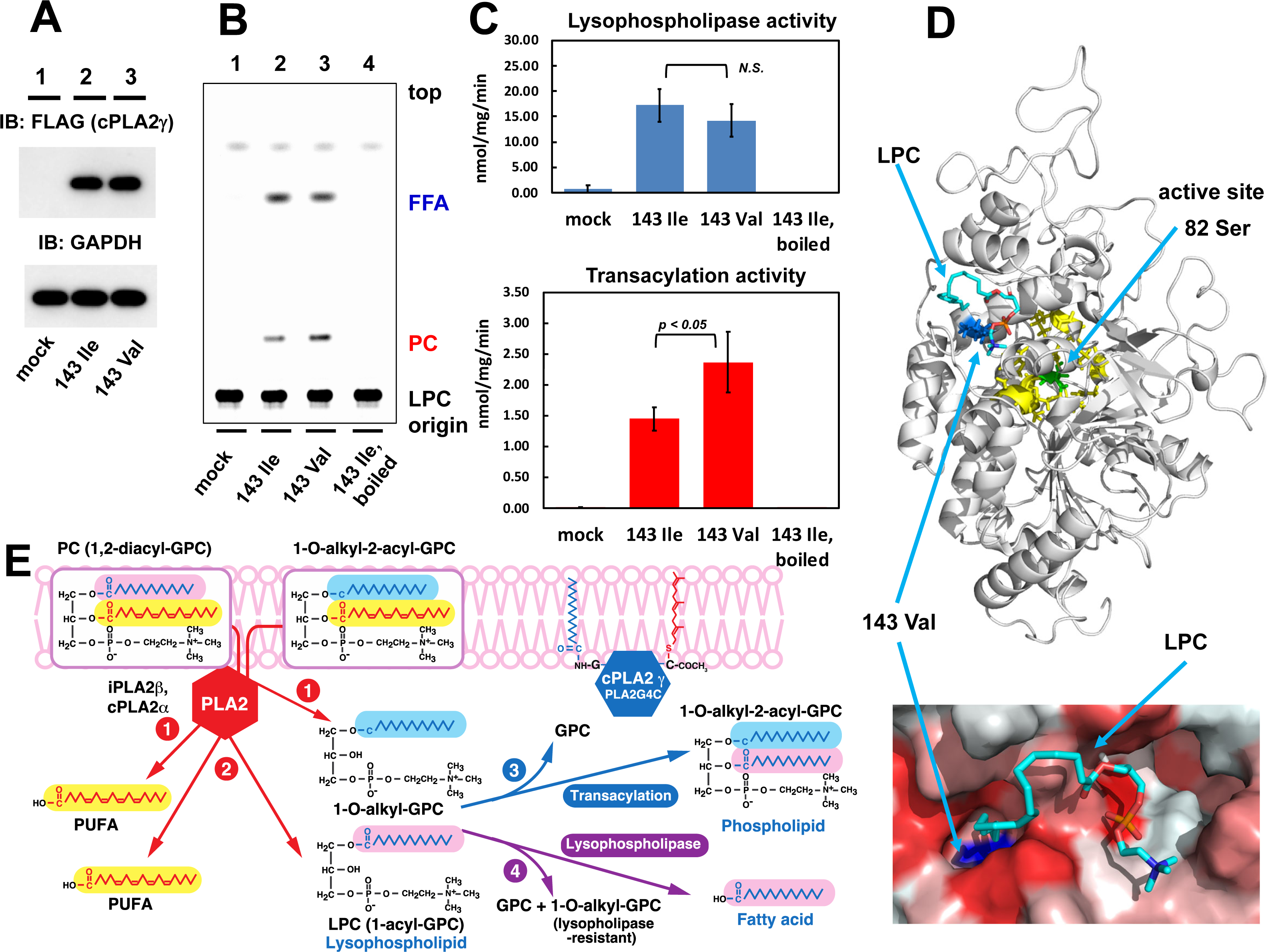
**Lysophopholipase and transacylation activities of transiently expressed cPLA2γ with Ile143 and Val143.** A: Expression of cPLA2γ with 143Ile and 143Val in HEK293 cells. The empty vector (mock, lane 1), pcDNA4/TO-FLAG-cPLA2γ with 143Ile (lane 2) or 143Val (lane 3) were transfected to HEK293 cells, and the expression of the recombinant proteins was evaluated by western blotting with anti-FLAG antibody (*upper*). Western blotting was also performed with anti-GAPDH antibody as a loading control (*lower*). The blot is a representative and similar results were obtained by three independent gene-transfections in HEK293 cells. IB = immunoblotting. B: TLC analysis of reaction products. The substrate [^14^C]LPC was incubated with the cell extract from mock (lane 1), FLAG-cPLA2γ-Ile143 (lane 2, MSA risk allele), or cPLA2γ-Val143 (lane 3)-transfected HEK293 cells at 37°C for 10 min. As a control, the substrate [^14^C]LPC was also incubated with the cell extract from FLAG-cPLA2γ-Ile143-transfected cells after inactivation by boiling (lanes 4). The substrate and products were extracted and separated by TLC, and quantified by Typhoon FLA9000 imaging. The TLC image is a representative and similar results were obtained in duplicate experiments with three independent gene-transfections in HEK293 cells. C: Lysophospholipase (*upper*) and transacylation (*lower*) activities were determined by the quantification of the radioactivity in FFA and PC, respectively. Values represent the mean ± S.D. from three independent gene-transfected cell experiments. *N.S.*, not significant. D: Consideration of the spatial position relationship between the Val143 and active Ser82 residues in cPLA2γ. The 3D-structure of cPLA2γ is predicted by the SWISS-Model and docking of cPLA2γ and LPC was analyzed using Autodock vina. The whole view (*upper*), and the surface view of the hydrophobic cavity near Val143 (*lower*) are shown. The active Ser82 and Val143 are shown by green and blue, respectively. The corresponding amino acids involved in the catalytic action of cPLA2α ^30^ are shown in yellow (the amino acid residues of Val81, Gly83, Ser84, Thr85, Trp86, Gly240, Ala242, Leu243, Asp385, Gly387, and Asn391). LPC (1-palmitoyl-*sn*-glycero-3-phospohocholine) is superimposed in the hydrophobic cavity near Val 143 (Ile143). The hydrocarbons of fatty acid, glycerol, and choline are shown in cyan, and oxygen and nitrogen atoms of glycerol and choline are in red and blue, respectively. In the *lower* panel, the hydrophobicity gradients are shown in red to white. The location of Val143 (or Ile143) is predicted in the surface of hydrophobic cavity in the opposite side of the catalytic pocket including Ser82 residue. In the *upper* panel, the catalytic domain including Ser82 residue faces the opposite direction from the viewer, while the hydrophobic cavity including Val143 (or Ile143) faces to the viewer. The distance between Ser82 and Val143 (Ile143) is calculated as approximately 15.8 A. The docking of LPC to the surface in the hydrophobic cavity is also predicted; the fatty acid and choline of LPC face hydrophobic and hydrophilic residues, respectively, and the location of the omega carbon of the fatty acid is very close to Val143 (or Ile143). This LPC may serve as acyl acceptor in the transacylation reaction. We propose that the amino acid substitution from Ile to Val increases the hydrophobic pocket and may result in the augmented transacylation activity by increased affinity for the acceptor LPC. E: Proposed roles of cPLA2γ in the remodeling of phospholipids. The fatty acid remodeling involving PLA2s (iPLA2β and cPLA2α) and cPLA2γ is depicted. The cPLA2γ hydrolyzes PC (1, 2-diacyl-GPC)) or 1-*O*-alkyl-2-acyl-GPC at the *sn*-2 position and releases polyunsaturated fatty acid (PUFA) and LPC (1-acyl-GPC) or 1-*O*-alkyl-GPC (reactions 1 and 2 in red). LPC and 1-*O*-alkyl-GPC metabolize the transacylation activity of cPLA2γ (reaction 3 in blue) through direct transfer of the *sn*-1 fatty acid of LPC to the *sn*-2 position of 1-*O*-alkyl-GPC to form 1-*O*-alkyl-2-acyl-GPC. The lysophospholipase activity (reaction 4 in magenta) of cPLA2γ hydrolyzes LPC at the *sn*-1 position to release fatty acid and GPC. The transacylation reaction is advantageous for remodeling 1-*O*-alkyl lysophospoholipids which are resistant to lysophospholipase. The transacylation reaction has also more advantageous than remodeling through acyl-CoA synthetase and acyltransferases, because acyl-CoA synthetase needs energy (ATP) consumption for activation of free fatty acid. In addition, the activity of acyl-CoA acyltransferase for 1-*O*-alkyl lysophospoholipids is much lower than that for 1-acyl lysophospoholipids ^39^.

In contrast, no reaction products were observed using mock (empty pcDNA4/TO vector)-transfected cell lysate. Recombinant FLAG-cPLA2γ catalyzes lysophospholipase activity but as shown by the release of [^14^C]FFA from [^14^C]LPC, the recombinant enzyme also catalyzes the transacylation reaction of [^14^C]FFA from [^14^C]LPC to another [^14^C]LPC to form [^14^C]PC (**Fig. 4B, E**).

Next, the activities of alternate alleles, cPLA2γ-Ile143 and cPLA2γ-Val143, were compared. Although the lysophospholipase activities for both enzymes were similar, the transacylation activity of cPLA2γ-143Val was significantly higher than that of cPLA2γ-Ile143 (*P* = 0.038) (**Fig. 4B, C**). We conclude the cPLA2γ p. Ile143Val amino acid substitution alters the transacylation activity of cPLA2γ.

The three-dimensional (3D) structural relationship between Ile143 and the active Ser82 residue of cPLA2γ was predicted by SWISS-Model based on the 3D-structure of cPLA2α and cPLA2δ, extrapolated from X-ray crystallography data (**Fig. 4D**) ^30, 31^. *In silico* docking experiments were carried out between the modelled 3D-structure of cPLA2γ and LPC (1-palmitoyl-*sn*-glycero-3-phosphocholine) using Autodock Vina^32^. The location of Ile143 (Val143) was predicted in a surface hydrophobic cavity opposite the catalytic pocket including Ser82; the distance between Ser82 and Ile143 was calculated as approximately 15.8 Å. Docking analysis revealed that LPC could bind the hydrophobic cavity near Ile143 (Val143) (**Fig. 4D**).

This LPC may serve as acyl acceptor in the transacylation reaction. However, the Ile143 to Val sidechain substitution results in a cavity that is larger by one methyl unit, which conceivably diminishes its transacylation activity (**Fig. 4D, E**).

## DISCUSSION

Genome-wide association analysis of MSA in Japanese population and subsequent meta-analysis in Japanese, Korean and Chinese patients demonstrates that rs2303744 G>A is significantly associated with MSA (*P* = 5.0 × 10^-15^ **Fig. 2B**, **Table 2**). In East Asia, the rs2303744 ‘A’ risk allele is marginally more prominent in the cerebellar subtype (MSA-C) (*P* = 8.6 × 10^-14^. OR = 1.57; 95% CI, 1.39 to 1.77) than in the parkinsonism subtype (MSA-P) (*P* = 7.2 × 10^-6^. OR = 1.38; 95% CI, 1.20 to 1.59) and the AA genotype is associated with younger onset disease (*P* = 0.001). While the association of rs2303744 with MSA was supportive in North American and European populations, it was considerably less significant than in the Japanese population and East Asia. Differences in the effect sizes observed may reflect, in part, the frequencies of the rs2303744 ‘A’ risk allele which is 0.39–0.46 in East Asia (Japan, Korea and China) compared to 0.77–0.78 in European/North American populations (**Table 2; Supplementary Fig. 3**). However, they may also reflect population differences in rs2303744 tagged haplotype composition and linkage disequilibrium. The high frequency of the rs2303744 ‘A’ risk allele and modest effect size, most especially in MSA in North American and European populations, raises a possibility of variability in *cis* that may contribute to or underlie the association. Further analysis of rs2303744 ‘A’ risk alleles and associated haplotypes by long-read sequencing is warranted in MSA cases.

The rs2303744 SNP, located in exon 5 of *PLA2G4C* c.427G>A, encodes p.Val143Ile allelic isoforms of cytosolic phospholipase A2 gamma (cPLA2γ). cPLA2γ was originally identified as a phospholipase A2 (PLA2) belonging to the *PLA2G4* family^33^. It is predominantly expressed in the brain, heart and skeletal muscle^34, 35^ and appears to be involved in remodeling and clearance of membrane phospholipids^36–38^. cPLA2γ has two enzyme activities catalyzing both lysophopholipase and transacylation reactions. Curiously, while lysophospholiapse activity is equivalent for cPLA2γ Val143 and Ile143 isoforms, we demonstrate cPLA2γ Ile143 has decreased transacylation activity (**Fig. 4B, C, E**). The physiologic consequence, if any, is unclear. However, activation of PLA2 enzymes, including cPLA2 (PLA2G4A) and iPLA2 (PLA2G6A), will lead to lysophospholipid accumulation if those products are not reacylated through acyl-CoA acyltransferases^39^. High levels of 1-*O*-alkyl or 1-*O*-alkenyl phospholipids are present in brain^34^, and 1-*O*-alkyl or 1-*O*-alkenyl lysophospholipids are formed primarily by the activation of PLA2. However, acyl-CoA acyltransferase activity for 1-*O*-alkyl or 1-*O*-alkenyl lysophospholipids is lower than that for 1-acyl lysophospholipids^39^. In addition, 1-*O*-alkyl or 1-*O*-alkenyl lysophospholipids are resistant to degradation by lysophospholipase activity. The transacylation activity of cPLA2γ is able to remodel the ether-linked lysophospholipids to phospholipids through the transfer of fatty acid of sn-1 position of 1-*O*-acyl lysophospholipids to ether-linked lysophospholipids (**Fig. 4E**). Hence, the higher clearance of *sn*-1 ether-linked lysophospholipids by cPLA2γ-Val143 over cPLA2γ-Ile143 may also play a protective role against neurodegeneration.

Intriguingly, the physiologic and pathologic roles of α-synuclein have been shown to be intimately related to lipid composition and trafficking^40–43^. Of interest, recessively-inherited mutations in iPLA2 (*PLA2G6*) cause infantile neuroaxonal dystrophy (neurodegeneration with brain iron accumulation-2A) with clinical presentations of dystonia/parkinsonism and cerebellar ataxia^44^. Pathologic features include prominent Lewy bodies/neurites (albeit without notable GCIs), neurofibrillary tangles/threads and axonal spheroids, and often associated with iron deposition in the globus pallidus and substantia nigra^45^. iPLA2 (*PLA2G6*) is also involved in remodeling phospholipids through the release of sn-2 fatty acid and 1-radyl lysophospholipid. Although no pathology or α-synuclein biology in the context of cPLA2γ has been examined, the present study suggests altered cPLA2γ expression and/or transacylase activity may underlie remodeling of membrane phospholipids and α-synuclein aggregation in oligodendroglia. Further genomic and functional studies are necessary to elaborate the role of cPLA2γ in neurons versus oligodendroglia, its effect on α-synuclein biology, and the pathobiologic mechanism of this devastating neurodegenerative disease.

## METHODS

### Subjects and DNA extraction

Demographic data of the participants enrolled in this study were summarized in Table 1. The MSA cases have been recruited by the Japan Multiple System Atrophy Consortium (JAMSAC). The diagnosis of MSA was made on the basis of the consensus criteria^3^. Pathologically confirmed MSA cases (definite MSA) are not included. The Japanese series (GWAS stage) comprised of 382 MSA cases and 388 control participants (the Japanese GWAS series). Pathologic expansions of trinucleotide repeats in *DRPLA*, *SCA1*, *SCA2*, *MJD/SCA3*, *SCA6*, *SCA7*, *SCA12*, and *SCA17* were examined and found to be in the normal range. Replication studies were conducted on a second Japanese series comprised of 373 original MSA cases and 495 control participants with no overlaps with the Japanese GWAS series. The Korean series comprised of 642 MSA cases and 446 control participants, the Chinese series comprised of 202 MSA cases and 274 control participants, the European series comprised of 154 MSA patients and 162 control participants, and the North American series comprised of 172 MSA cases and 294 control participants^27^. Genomic DNA was isolated from patient’s blood leukocytes using standard procedures. The research project was approved by Research Ethics Committee of the Faculty of Medicine of the University of Tokyo. The research projects were approved by the participating institutional review boards. Written informed consent was obtained from all the human research participants.

### SNP genotyping at GWAS stage

SNP genotyping was conducted in Japanese series comprising of 382 cases and 388 controls (GWAS stage) using HumanHap 550 and Human660W-Quad BeadChips (Illumina) according to the manufacturer’s instructions. Genotypes were called and extracted using BeadStudio version 3.1.3.0 (Illumina). The average overall call rate was 99.7%. The principle component analysis (PCA), along with HapMap data (CEU, YRI, and JPT+CHB) using EIGENSTRAT^46^, confirmed that there are no outliers suggesting different ethnicity. Quality control analyses were conducted using PLINK 1.9 0b6^47^. Three controls with a pi-hat > 0.2 were excluded from the analysis. None of the individuals were excluded with more than 2% missing genotypes. As the result, 382 cases and 385 controls were included in the following analysis. SNPs were excluded according to the following criteria: (1) missing call rates > 0.01, (2) minor allele frequency < 0.05, (3) Hardy–Weinberg equilibrium (HWE) *P* < 0.01, and (4) SNPs not on the 22 autosomes. A total of 456,818 informative SNPs were included in the GWAS. A Manhattan plot and quantile-quantile plot were constructed using the R 4.0.1. Imputation analysis was performed with IMPUTE4 to investigate associations of ungenotyped SNPs^48^. We used the Japanese reference provided by the Tohoku University Tohoku Medical Megabank Organization (ToMMo) and the following criteria were employed; i) probability of best guess genotype >0.9 and ii) Information Score >0.4. We defined empirical genome-wide significance as *P* < 1.09 ×10^-7^ and evidence for suggestive association as *P* < 1.0×10^-6^.

### Replication study

For replication studies, the SNP (rs2303744) identified at the GWAS stage was analyzed in the Japanese (replication), Korean, Chinese, European and North American series. For the Japanese series (replication) of MSA cases and controls, we utilized sequence data of rs2303744 obtained from exome analysis. The genomic DNA was captured using SureSelect v4+UTRs or SureSelect v5+UTRs. Exome sequence analysis was conducted using Illumina Hiseq 2500 (83-bp or 101-bp paired-end). BWA^49^ and Samtools^50^ were used with the default parameter settings for alignment and variant detection against the human reference genome (NCBI37/hg19). Genotyping for rs2303744 (an average sequencing depth of 58.5×) required a minimum of reads of 8× or greater in all participants in the Japanese exome series. For the Korean series, genotyping was performed by TaqMan SNP genotyping (Thermo Fisher Scientific). For the Chinese series, genotyping of rs2303744 was achieved by Sanger sequencing. For the European and North American series, we utilized SNP data of rs2303744 obtained from SNP typing using Human660W-Quad BeadChips (Illumina) as described above.

### Statistical analysis

The associations between SNPs and MSA were assessed by chi-squared test with a two-by-two contingency table in an allele frequency model using PLINK. In the replication studies, chi-squared test was used to test the associations of rs2303744 with MSA in individual series using R 4.0.1. Regional association plots were produced with LocusZoom (v1.3) (http://locuszoom.org/). The meta-analysis was conducted using the random effects model (DerSimonian-Laird method) or the fixed-effects model (Mantel-Haenszel method) to calculate pooled odds ratio in accordance with Cochran’s Q test as a measure of heterogeneity^51^. Multiple logistic regression analysis and multiple linear regression analyses were conducted using the glm() function in R 4.0.1.

### Construction of Plasmids and Mutagenesis

cDNA clone of Phospholipase A2 group IVC (cPLA2γ) was purchased from Kazusa DNA Research Institute (Clone No.FXC26716E). FLAG-tag fused full-length cPLA2γ expression plasmids (FLAG-cPLA2γ:pcDNA4/TO) were generated by subcloning the PCR products into the KpnI/NotI site of pcDNA4/TO (Thermo Fisher:V102020) FLAG-fused forward primer: cc ggt acc gcc gcc ATG GAC TAC AAA GAC GAT GAC GAC AAG gga ttc GGA AGC TCT GAA GTT TCC ATA. Reverse primer:tt gcggccgc CTA TGC CAA GCA GCA ACT TCG GGC (cording sequence shows a capital letter). FLAG-cPLA2γ-143Val (FLAG-cPLA2γ-I143V:pcDNA4/TO) were obtained by the mutagenesis of FLAG-PLA2G4C:pcDNA4/TO using KOD -Plus-Mutagenesis Kit (TOYOBO:SMK-101). Upper primer (mutation shown as a capital letter); atggttGtctctaagcaaaccagaga. Lower primer: gtaggcccagaagtcggtca. All constructs were verified by nucleotide sequence analysis (ABI PRISM 3100 Genetic Analyzer, Applied Biosystems) with BigDye terminator ver3.1 (Life Technologies 4337455). All vectors were verified for expression by Western blot.

### Functional analysis of *PLA2G4C* variants

HEK293 cells (4 × 10^5^ cells) were plated in 6-well plates and cultured with DMEM supplemented with 10% FBS, 100 µg/mL streptomycin, and 100 U/mL penicillin for 24h, and the cells were transfected with 1 µg of plasmid using Lipofectamine 2000; The plasmids were pcDNA4/TO (control), pcDNA4/TO/FLAG-cPLA2γ-143Val, and pcDNA4/TO/FLAG-cPLA2γ-143Ile (risk allele). Six independent transfections were done for each plasmid. After transfection, the cells were cultured for an additional 24 h and the cell extract was prepared. The cells were sonicated in buffer containing 50 mM Tris-HCl (pH 7.5), 2 mM EDTA, complete protease inhibitor (Roche) using a bath-type sonicator.

The expression of each recombinant protein was confirmed by Western blotting. The extracts from each cell were subjected to SDS-PAGE with WIDE-VIEW^TM^ Prestained Protein Size Marker III (Wako, Japan). After the proteins were transferred to a nitrocellulose membrane, the membrane was incubated with anti-FLAG antibody, followed by HRP-conjugated secondary antibody. The anti-GAPDH antibody was used for the loading control. Signals were detected with the Western BLoT Quant HRP substrate (Takara, Japan) and analyzed and quantified using the Amersham Imager 600 with ImageQuant TL software (GE Healthcare, USA).

Lysophospholipase/transacylation activity was determined by the measurement of [^14^C]oleic acid release or the formation of [^14^C]phosphatidylcholine (PC) from [^14^C]LPC (1-[^14^C]oleoyl-GPC). Aliquots of [^14^C]LPC (27.5 nCi/assay) were mixed with unlabeled LPC, and suspended in 50 mM Tris-HCl (pH 7.5) containing 2 mM EDTA. Each cell extract was incubated with 500 µM LPC in the assay solution at 37°C for 10 min, and the reactions were terminated by addition of chloroform:methanol (1:2, v/v). The cell extract (cPLA2γ-143Ile) after the inactivation by the boiling was also run in the same conditions as a control. Total lipids were extracted according to Bligh and Dyer. Total lipids were separated by silica gel TLC developed with chloroform:methanol:triethylamine: H_2_O (30:35:35:7, v/v/v/v).

Radioactive bands corresponding to substrate (LPC) and products (FFA and PC) were determined in an Typhoon FLA9500 (GE Healthcare, USA). The activity was calculated as the release of [^14^C]oleic acid (lysophospholipase) or the formation of [^14^C] PC (transacylation activity). The results are shown as the mean ± S.D. in duplicate enzyme reactions with three independent transfected cells. For comparisons, the Student’s t-test (both sided) were performed after equal variance test was confirmed with GraphPad PRISM 6 (GraphPad Software, USA).

### Prediction of 3D structure *and* LPC-binding mode of *PLA2G4C*

The 3D-structure of cPLA2γ is predicted by SWISS-Model by using the top-ranked homologous structures as the templates of homology modeling. The docking of LPC (1-palmitoyl-*sn*-glycero-3-phospohocholine) into the modelled 3D-structure of cPLA2γ were carried out by using Autodock vina. The grid centers for the Vina docking were selected at two sites, the active Ser82 and the Ile/Val143, with the dimensions 40 Å × 40 Å × 40 Å.

## Data Availability

The data that support the findings of this study are available from the corresponding author upon reasonable request.

## Acknowledgments

We deeply thank the patients for participating in the study. We acknowledge Drs. Takahisa Kawaguchi and Tatsuhiko Tsunoda at RIKEN Center for Integrative Medical Sciences, Yokohama, Japan, for their kind advices for the genome-wide association study including the PCA analysis. We also acknowledge Prof. Yukinori Okada for his valuable advice on the effect of regional differences in the genome variations on the results of GWAS. This work was supported in part by KAKENHI (Grants-in-Aid for Scientific Research on Innovative Areas Nos. 22129001 and 22129002) from the Ministry of Education, Culture, Sports, Science and Technology of Japan, Grants-in-Aid [H23-Jitsuyoka (Nanbyo)-Ippan-004 and H26-Jitsuyoka (Nanbyo)-Ippan-080] from the Ministry of Health, Welfare and Labour, Japan, and grants (15ek0108065h0002, 16kk0205001h0001, 17kk0205001h0002, 17ek0109279h0001, 18ek0109279h0002, 19ek0109279h0003, 20ek0109491h0001, 21ek0109491h0002, and 22ek0109491h0003) from the Japan Agency for Medical Research and Development (AMED) to S.T. This work was in part supported under the framework of international cooperation program managed by National Research Foundation of Korea (Grant Number: NRF-2015K2A1A2070274) to B. J. Korean DNA samples for this study were provided by the Seoul National University Hospital Human Biobank, a member of the National Biobank of Korea, which is supported by the Ministry of Health and Welfare. All samples derived from the National Biobank of Korea were obtained with informed consent under institutional review board-approved protocols and we thank Erika Scott for technical assistance. S.W.S. was supported by the Intramural Research Program of the National Institute of Neurological Disorders and Stroke, U.S. National Institutes of Health (program #: 1ZIANS003154).

## Competing Interest Statement

S.T. received a research grant from Nobelpharma Co., Ltd. S.W.S. serves on the Scientific Advisory Council of the Lewy Body Dementia Association and the Multiple System Atrophy Coalition. S.W.S. receives research support from Cerevel Therapeutics. S.W.S. is an editorial board member of the Journal of Parkinson’s Disease and JAMA Neurology.

## Notes

### Author Declarations

The research project was approved by Research Ethics Committee of the Faculty of Medicine of the University of Tokyo.

