## Supplementary Table 1, Table 2, Supplementary Fig.1, Fig. 2, Fig. 3 for "Genome-wide association study identifies a new susceptibility locus in *PLA2G4C* for Multiple System Atrophy": SupplementaryMaterial_08.pdf

**Supplementary Table 1. Allelic association test of rs2303744 (PLA2G4C) with Japanese multiple system atrophy (MSA) cases in combined stage with reference to Controls in 38K JPN**

| dbSNP rs ID | Gene | Risk Allele | Allele | Case | | Control | | OR | *P*-Value |
|---|---|---|---|---|---|---|---|---|---|
| | | | | A | G | A | G | | |
| rs2303744 | *PLA2G4C* | A | A/G | 771 (0.511) | 739 (0.489) | 30297 (0.3912)* | 47147 (0.6088)* | 1.62 (1.46-1.80) | $2.2 \times 10^{-16}$ |

* Controls in 38K JPN (Integrative Japanese Variation Database, Tohoku University Tohoku Medical Megabank Organization (ToMMo) (https://jmorp.megabank.tohoku.ac.jp/). Odds ratio (OR) of the effect with respect to the allele A and association P-values are given.

**Supplementary Table 2. Multiple logistic regression analysis of rs2303744 (PLA2G4C) genotypes and MSA-C and MSA-P in East Asian populations (adjusted for sex and population)**

| | Odds ratios | 95% Confidence interval | P value |
|---|---|---|---|
| MSA-C | 1.57 | 1.39 – 1.77 | $8.6 \times 10^{-14}$ |
| MSA-P | 1.38 | 1.20 – 1.59 | $7.2 \times 10^{-6}$ |

Number of cases and controls

MSA-C: 546 (Japanese), 262 (Korean) and 121 (Chinese)
MSA-P: 181 (Japanese), 320 (Korean) and 51 (Chinese)
Controls: 880 (Japanese), 446 (Korean) and 274 (Chinese)

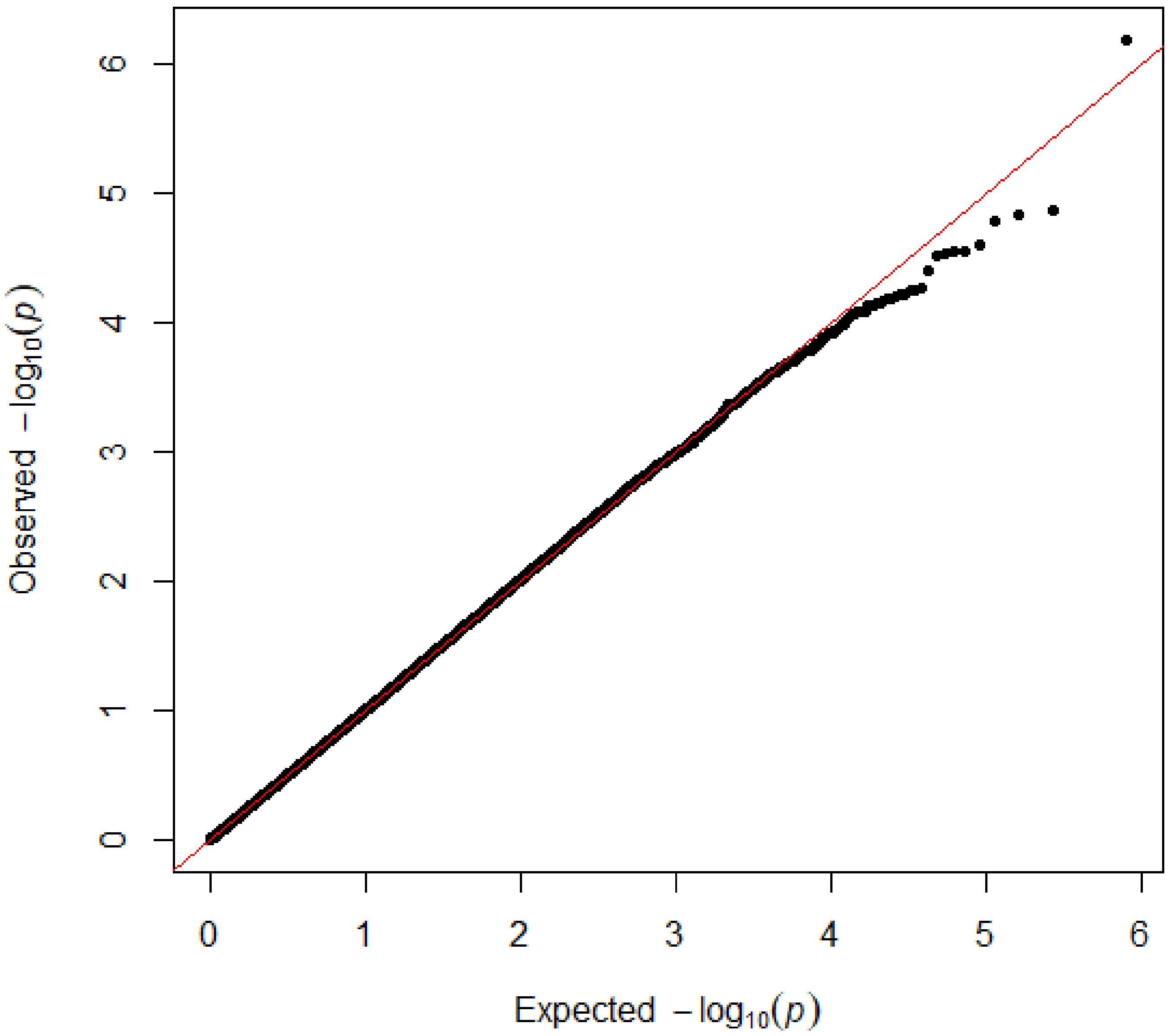


**Supplementary Fig. 1** Quantile-quantile plots (QQ-plots) of the genome-wide association study.

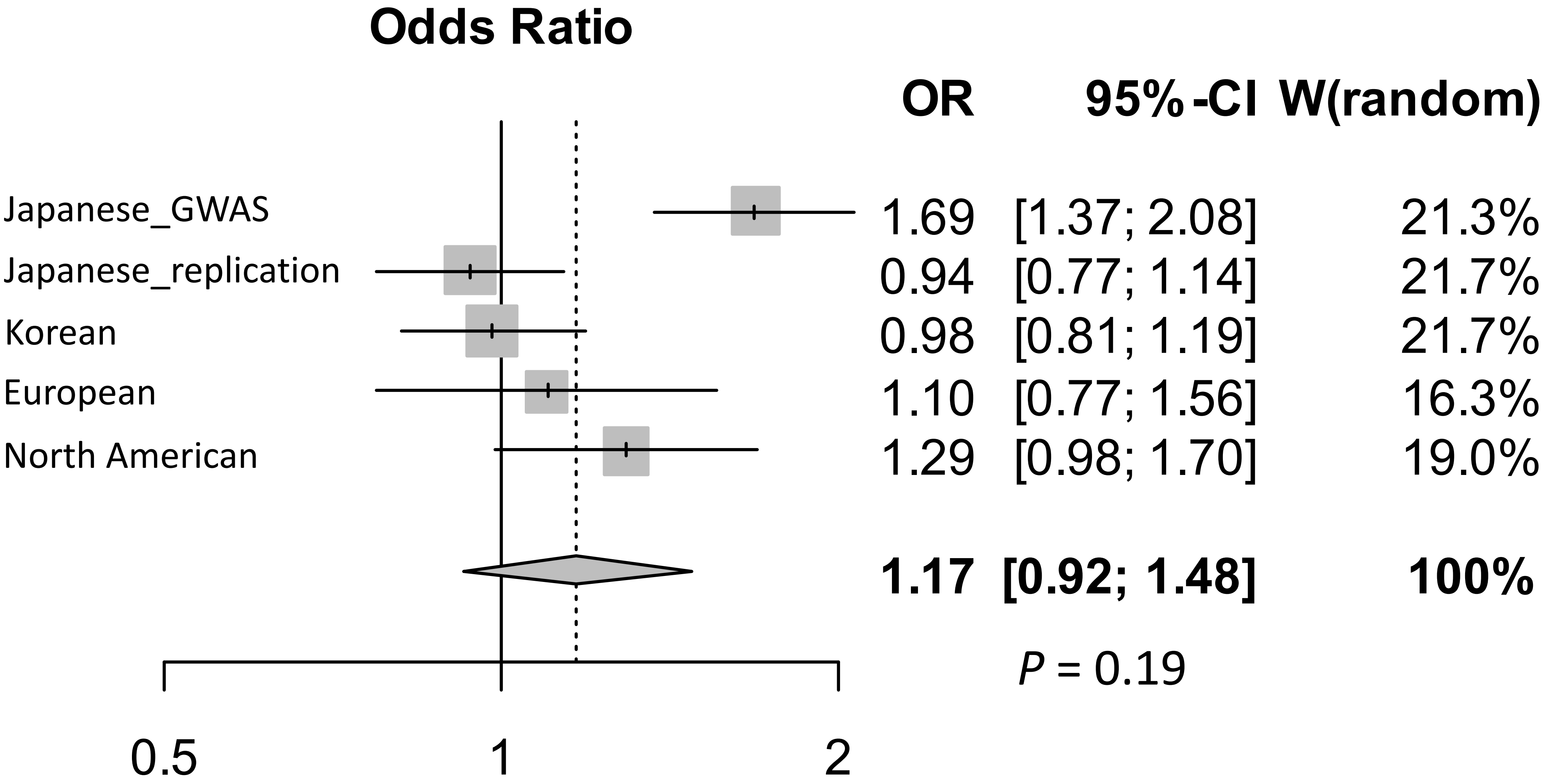


**Supplementary Fig. 2 Forest plots for showing the meta-analysis of rs1569325.**
Squares and horizontal lines represent estimated ORs and 95% CIs for individual series. The size of each square represents the statistical weight, the inverse of the variance, in each study. A diamond shows the summary OR estimate and 95% CIs for the meta-analysis. The area of diamond is proportional to the sum of the logarithm of odds ratios weighted by the inverse of the variance of each study.

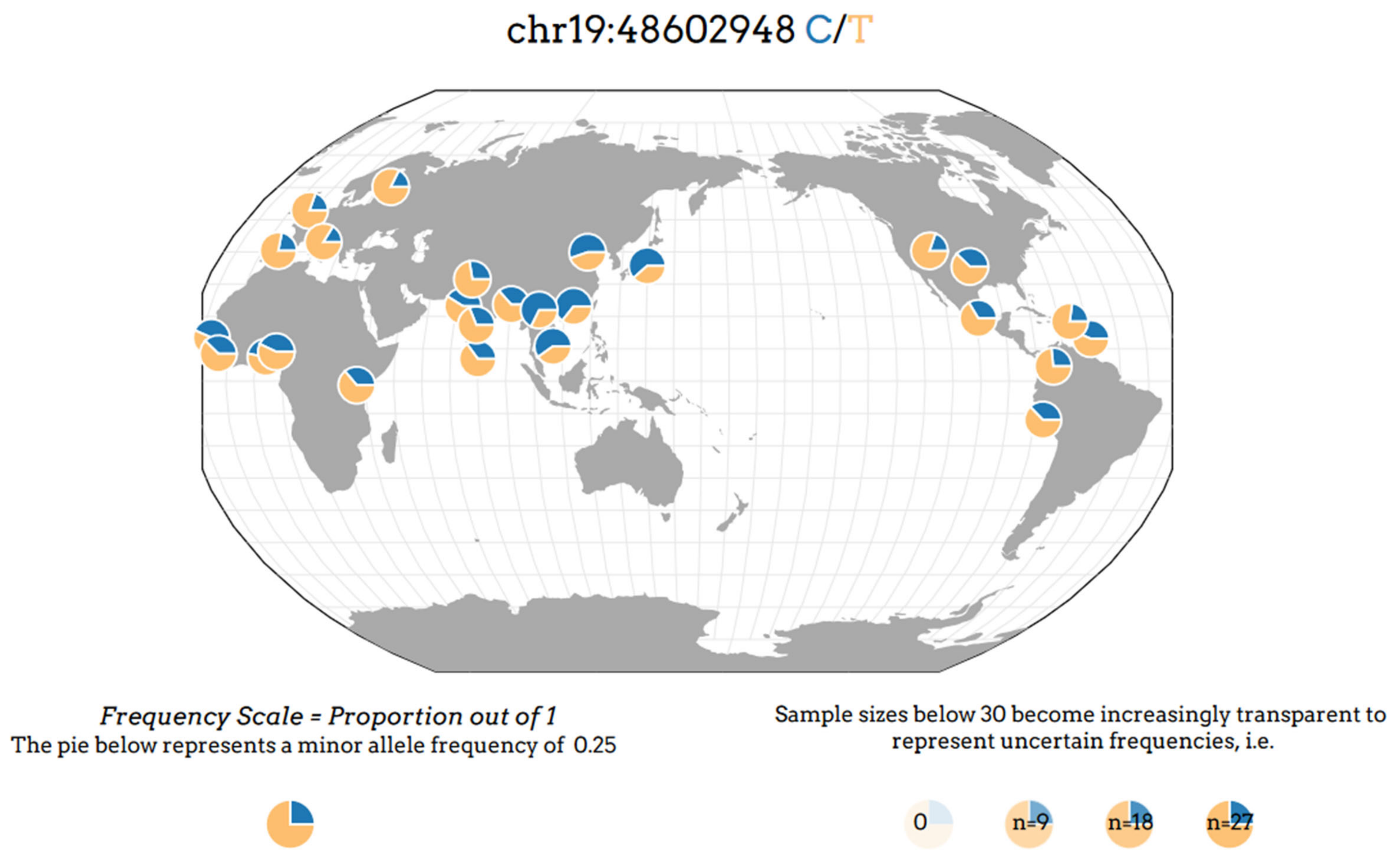


**Supplementary Fig. 3 Geographical distribution of T/C alleles (rs2303744).**
The geographical distribution of the T/C alleles of rs2303744 was generated using the Geography of Genetic Variants Browser (https://popgen.uchicago.edu/ggv/). The T allele is the major allele in the European and North American populations, while allele frequencies of the C allele are much larger in East and South Asian populations than those of European and North American populations. The map position of rs2303744 is based on GRCh19/hg19.
